# Factors Associated with Timely Test Seeking, Test Turnaround, and Public Reporting of COVID-19: a retrospective analysis in Ontario, Canada

**DOI:** 10.1101/2021.02.22.21252219

**Authors:** Eugene Joh, Sarah A. Buchan, Nick Daneman, Lauren A. Paul, Kevin A. Brown

## Abstract

**Background:** Minimizing delays in disease identification and reporting improves the timeliness of surveillance data, and can reduce transmission of COVID-19. Our study investigates factors associated with timely testing and reporting of COVID-19 during the first pandemic wave in one province of Canada.

**Methods:** We identified all persons with confirmed SARS-CoV-2 infection residing in private households across the largest province of Canada, Ontario from the date of the first confirmed case in Ontario (January 25) to July 19, 2020. Our primary outcomes consisted of: (1) specimen collection within 1 day of symptom onset (test seeking), (2) test result reported to local public health within 1 day of specimen collection (test turnaround), and (3) entry of case data into the provincial database within 1 day of reporting test results (reporting). We examined 14 covariates including eight case characteristics, and six neighborhood characteristics. In addition to descriptive measures, logistic regression models were fitted. Unadjusted models included the covariate alone, while adjusted models included age, gender, month, and region.

**Findings:** Among 27,198 COVID-19 cases from January 25 2020 to July 19 2020, 28·7% had timely test seeking, 40·2% had timely test turnaround, and 75·5% had timely reporting. Male gender had lower odds of timely test seeking (adjusted odds ratio [aOR] 0·79 [95% CI: 0·74-0·85]) compared to females. Healthcare worker status (aOR 2·77 [95% CI: 2·52-3·05] compared to non-healthcare workers), and age ≥80 years (aOR 1·59 [95% CI: 1·33-1·91] compared to 40-59 year olds) were associated with timely test seeking. Specimen collection on Fridays and Saturdays (aOR 0·88 [95% CI: 0·79-0·98], aOR 0·83 [95% CI: 0·74-0·92] respectively, compared to Wednesdays) had lower odds of timely test turnaround. Urban areas (aOR 1·55 [95% CI: 1·41-1·70] compared to rural areas) were associated with timely test turnaround. Urban areas (aOR 0·79 [95% CI: 0·70-0·89] compared to rural areas) were less likely to have timely reporting.

**Interpretation:** Individual, neighborhood, and administrative factors are associated with timely testing and reporting of SARS-CoV-2 infections. These findings present considerations for developing targeted strategies to minimize delays and improve timely testing and reporting of SARS-CoV-2 infections.

**Funding:** This study was funded by Public Health Ontario.

**Research in Context:** *Evidence before this study:* We searched PubMed and medRxiv up to November 30 2020 to identify studies examining the impact of delays in the disease reporting process on the public health response to COVID-19. We used the search terms (“2019-nCoV” OR “COVID-19” OR “SARS-CoV-2”) AND (“delays” OR “timely” OR “reporting” OR “test” OR “turnaround”), and reviewed reference lists of any relevant articles in the original search. Numerous modeling studies have highlighted the importance of timely testing and reporting to effectively control the spread of COVID-19. Additional studies have also identified delays of only 1 day in testing were associated with increased risk of secondary transmission within households. However no study has described the multiple delays in the disease reporting process of COVID-19 and examined factors associated with timely disease reporting using a large population cohort.

*Added value of this study:* Our study described timely test seeking, test turnaround, and reporting for laboratory-confirmed COVID-19 cases in Ontario, Canada and identified associated individual, neighbourhood, and administrative factors. To the best of our knowledge, this study is the first to describe detailed delays in the disease reporting process of COVID-19 and identified associated factors using a large population cohort.

*Implications of all the available evidence:* Numerous individual, neighborhood, and administrative characteristics are associated with timely testing and reporting of COVID-19. These identified factors may be used to develop strategies such as broadened test access, prioritization of vulnerable populations, and increased testing capacity to reduce delays in testing and reporting and improve the effectiveness of public health response to COVID-19.

## Introduction

Effective disease control via public health follow-up is contingent on rapid case identification to interrupt infection spread, and timely detection of secondary contacts. As case numbers increase, case and contact management may be delayed due to limited public health resources. Rapid case identification is critical to interrupting transmission, and the speed necessary to interrupt transmission varies across infections as a function of the time from symptom onset to the infectious period.^1^ Studies of COVID-19 viral kinetics and contact tracing have pointed to the prodromal phase of SARS-CoV-2 infection having the highest risk of transmission, with the peak of infectiousness thought to occur on and following the date of symptom onset, and culturable virus detected up to 9-10 days after symptom onset.^2–5^ This characteristic of COVID-19 points to the efficient spread of SARS-CoV-2 in the community and has made it challenging for public health to rapidly identify new cases before secondary transmission.

The process of disease reporting will inevitably have delays that can occur between symptom onset, specimen collection, laboratory testing, and reporting to public health. Minimizing both these individual and overall delays is critical in slowing the spread of COVID-19. Studies have shown longer delays are associated with increased risk of secondary household transmission and play a large role in whether COVID-19 outbreaks can be controlled.^6,7^

We sought to identify individual- and neighbourhood-level characteristics associated with timely testing and reporting of confirmed COVID-19 during the first COVID-19 wave in Ontario, Canada (defined as January to July 2020).

## Methods

### Study population

COVID-19 was designated as a mandatory reportable disease on January 22, 2020. From the provincial database we extracted information on persons with confirmed SARS-CoV-2 infection reported by laboratories to public health in Ontario, Canada from January 25, 2020 to July 19, 2020.^8^ As testing criteria, prioritization, and access differed across populations in Ontario, we excluded people belonging to congregate settings (e.g. nursing houses, retirement homes, shelters, agricultural settings, correctional facilities, and long-term care homes) based on criteria described elsewhere,^7^ thus restricting our cohort to residents of private households.

### Outcomes

We generated three timeliness metrics based on the delays in the disease reporting process for each individual with COVID-19. These metrics were defined as: (1) the time between a case’s symptom onset to when their specimen was collected for PCR testing (test seeking timeliness); (2) the time from when a case’s specimen was collected to when a positive test result was reported to local public health (test turnaround timeliness); and (3) the time between when the test result was reported to when local public health entered the case’s data in the provincial reportable disease systems (reporting timeliness). Our primary outcomes of interest were whether each metric was considered timely (delay of ≤ 1 day). The thresholds were based on previous evidence of increased odds of secondary household transmission even with 1-day delay of test seeking,^7^ and suggested limits in testing delays of successful contact tracing.^9,10^ Persons with symptom onset occurring after a positive PCR test were considered to have timely test seeking. Individuals were excluded when the date of reporting occurred before specimen collection, and when date of data entry occurred before date of reporting, as we were unable to determine whether these were legitimate entries or due to data entry error. Individuals with missing date information on symptom onset or specimen collection were excluded for the timely test seeking metric, and individuals with missing date information on specimen collection were excluded from the timely test turnaround metric. Information on the date of when a positive test result was reported to public health was complete for all individuals.

### Individual-level characteristics

Individual-level characteristics were extracted from the provincial reportable disease system: age, gender, and health region (Supplementary Material Table S1). Information on whether a case was part of a public health declared outbreak and whether the person’s occupation was identified as a healthcare worker (doctor, nurse, dentist, dental hygienist, midwife, other medical technicians, personal support worker, respiratory therapist, and first responder) were also included. We extracted temporal covariates including month and day of week for each timeliness metric.

Individuals were classified as either an index household case or a secondary household case based on symptom onset date (or specimen collection date when symptom onset information was not available). Index cases were defined as the first case identified within an individual household. Secondary cases of COVID-19 within households were categorized based on the number of days from the index case (1-5, 6-10, ≥11 days). Index and secondary cases linked to an outbreak were identified and categorized in the same manner (1-5, 6-10, ≥11 days from index case associated with an outbreak).

### Neighbourhood-level characteristics

Information for neighbourhood-level characteristics were obtained from 2016 Canadian census data based on dissemination areas.^11^ We linked individuals with COVID-19 to dissemination areas using postal codes; in rare instances when a resident postal code matched with multiple dissemination areas, the dissemination area with the largest population was selected. Neighborhood characteristics of interest included the percentage of non-White and non-Indigenous residents, recent immigration, wage below low-income cut-off (LICO), labour force status, attainment of high school education, and urban/rural status.

### Statistical Analysis

We performed descriptive analyses to assess the characteristics of timely cases. Chi-squared tests, Student’s t-tests, and Cochran-Mantel-Haenszel trend tests were used to test for differences in individual- and neighbourhood-level characteristics among people with timely and non-timely metrics. Distributions of the test seeking, test turnaround, and reporting delays were generated. We used logistic regression models to determine characteristics associated with timeliness for each of the three metrics. For each outcome we fit unadjusted and adjusted models that accounted for age, gender, month, and health region. We reported odds ratios, unadjusted (OR) and unadjusted (aOR), with 95% confidence intervals (95% CI) for each characteristic. Odds ratios for neighbourhood-level covariates were reported per 10% increases. All analyses were conducted using R version 3·6·3.

## Results

As of July 19, 2020, there were 27,198 cases of COVID-19 in Ontario occurring outside of congregate settings (Figure 1). Of these cases 10,295 (37·9%) had missing information on the date of symptom onset or date of testing, 6,525 (24·0%) were missing the date of testing, and no cases were missing the date test results were reported or the date of data entry. This resulted in 16,903 cases with complete test seeking delay information, 20,673 cases with test turnaround delay information and 27,198 cases with complete reporting information (Table 1).

**Table 1:** Timeliness of test seeking, test turnaround, and reporting.

|  | **Complete Cases**<br>**(N)** | **Timely**<br>**(N, %)** | **Duration of Delay (days)**<br>**Median (IQR)** |
| --- | --- | --- | --- |
| Test seeking | 16,903 | 4,859 (28.7%) | 3 (1, 7) |
| Test turnaround | 20,673 | 8,316 (40.2%) | 2 (1, 3) |
| Reporting | 27,198 | 20,535 (75.5%) | 1 (0, 1) |

**Figure 1:**
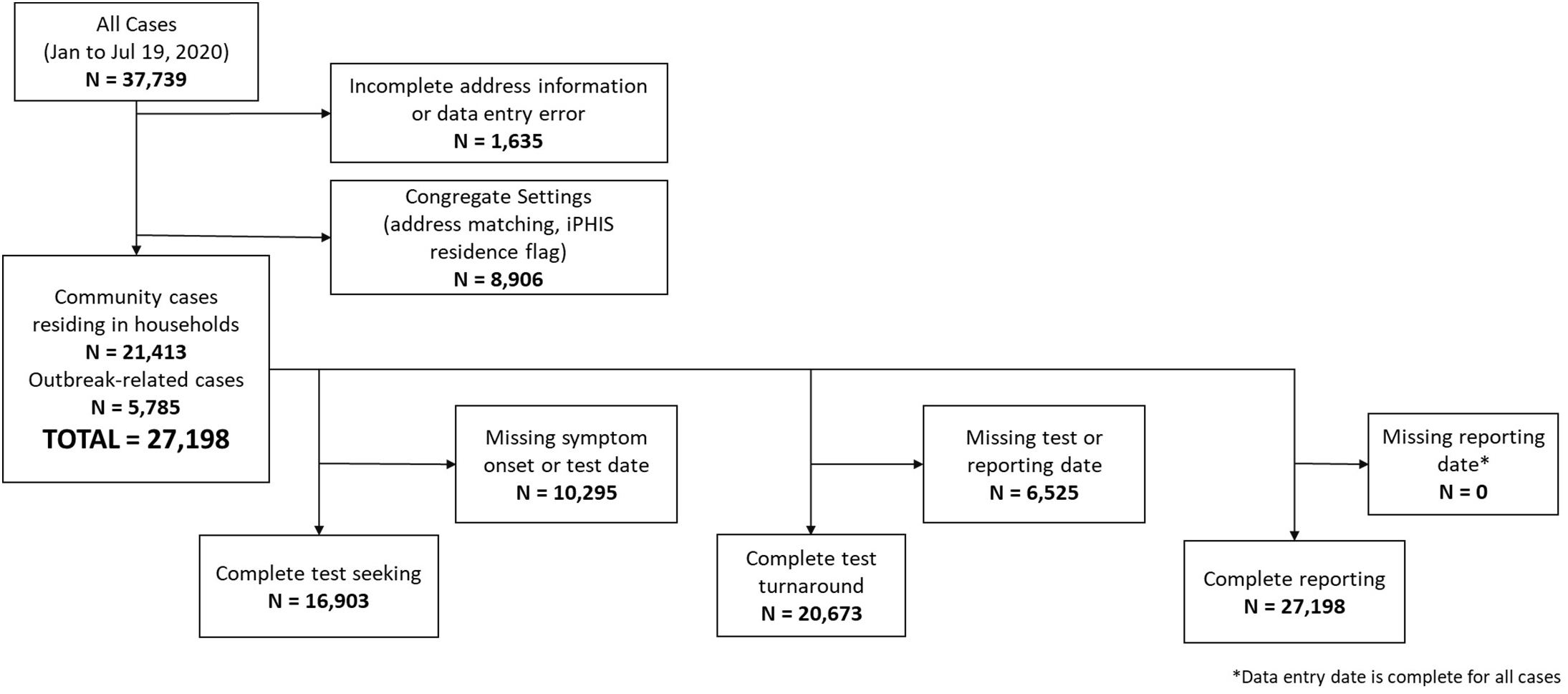
Cohort Flowchart

Across the three metrics, 28·7% of cases had timely test seeking, 40·2% had timely test turnaround, and 75·5% had timely reporting. Only 6·6% of individuals with COVID-19 were timely for all three metrics. The distribution of test seeking delays had greater dispersion (3 days, interquartile range [IQR]: 1-7) compared to test turnaround (2 days, IQR: 1-3) and reporting (1 day, IQR: 0-1) delays (Figure 2).

**Figure 2:**
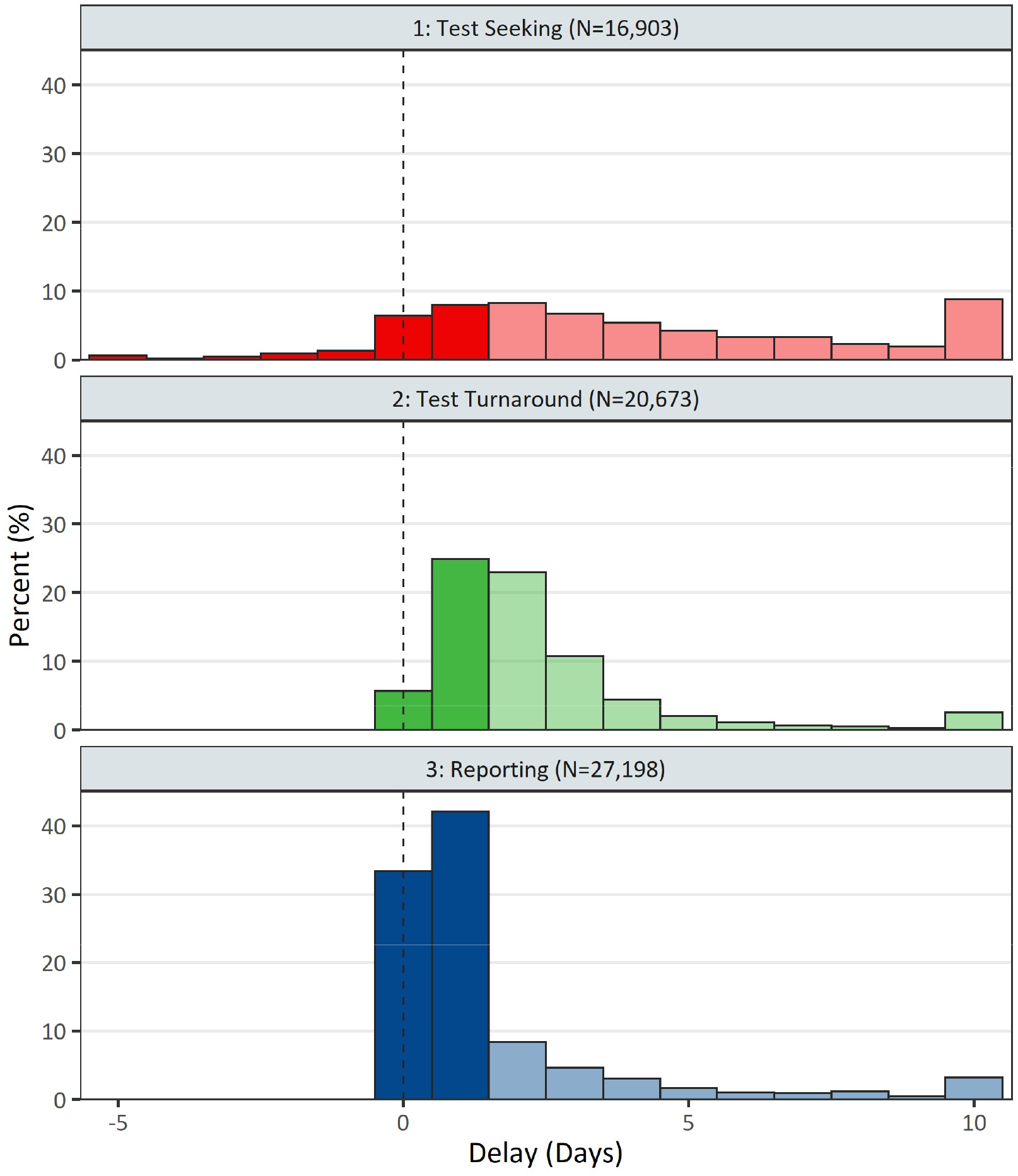
Distribution of Timeliness Metrics. Distribution of test seeking (symptom onset to specimen collection), test turnaround (specimen collection to test result), and reporting (test result to reporting) delays. Solid regions represent COVID-19 cases that were timely (≤1 day), while transparent regions represent cases that were delayed (>1 day). The dotted line denotes zero-day delay or events occurring on the same day.

### Timely Test Seeking

Males had significantly lower odds of timely testing compared to females (Table 2; aOR = 0·79 [95% CI: 0·74-0·85]). Healthcare workers (aOR = 2·77 [95% CI: 2·52-3·05]) and those aged ≥80 years (aOR = 1·59 [95% CI: 1·33-1·91]) had significantly higher odds of timely testing compared to cases with non-healthcare occupation and cases aged 40-59 years, respectively. Secondary cases in both outbreak and household settings were more likely to have timely testing compared to index cases. There was a trend of increasing odds of timely testing by month of symptom onset among cases from March (aOR = 0·29 [95% CI: 0·26-0·33]; reference = May 2020) to July (aOR = 2·17 [95% CI: 1·84-2·56]; reference = May 2020). Symptom onset occurring on days adjacent to or on a weekend were associated with lower odds of timely test seeking. Urban areas had lower odds of timely test seeking compared to rural areas. Neighbourhood factors were not strongly associated with test seeking.

**Table 2:** Factors associated with timely test seeking.

|  | **Total (N = 16,903)**<br>(Median [IQR] or<br>N[%]) | **Timely (N = 4,859)**<br>(Median [IQR] or<br>N[%]) | **p value** | **Odds Ratio**<br>(95% CI) | **Adjusted Odds**<br>**Ratio (95% CI)†** |
| --- | --- | --- | --- | --- | --- |
| **Month of Symptom Onset** |  |  | < 0·001 |  |  |
| March and Earlier | 3,818 | 483 (12·7%) |  | 0·30 (0·27 - 0·33) | 0·29 (0·26 - 0·33) |
| April | 5,704 | 1,675 (29·4%) |  | 0·85 (0·78 - 0·93) | 0·84 (0·77 - 0·91) |
| May | 4,576 | 1,499 (32·8%) |  | reference | reference |
| June | 2,138 | 860 (40·2%) |  | 1·38 (1·24 - 1·54) | 1·40 (1·26 - 1·56) |
| July | 667 | 342 (51·3%) |  | 2·16 (1·83 - 2·54) | 2·17 (1·84 - 2·56) |
| **Weekday of Symptom Onset** |  |  | 0·010 |  |  |
| Sunday | 2,207 | 618 (28·0%) |  | 0·91 (0·80 - 1·03) | 0·94 (0·83 - 1·07) |
| Monday | 2,712 | 787 (29·0%) |  | 0·96 (0·85 - 1·08) | 0·98 (0·87 - 1·11) |
| Tuesday | 2,427 | 742 (30·6%) |  | 1·03 (0·91 - 1·17) | 1·07 (0·95 - 1·22) |
| Wednesday | 2,405 | 720 (29·9%) |  | reference | reference |
| Thursday | 2,280 | 731 (32·1%) |  | 1·10 (0·98 - 1·25) | 1·10 (0·97 - 1·25) |
| Friday | 2,580 | 690 (26·7%) |  | 0·85 (0·76 - 0·97) | 0·85 (0·75 - 0·97) |
| Saturday | 2,292 | 571 (24·9%) |  | 0·78 (0·68 - 0·88) | 0·78 (0·69 - 0·89) |
| **Age (years)** |  |  | < 0·001 |  |  |
| 0-19 | 960 | 338 (35·2%) |  | 1·37 (1·19 - 1·58) | 1·10 (0·95 - 1·28) |
| 20-39 | 5,971 | 1,739 (29·1%) |  | 1·04 (0·96 - 1·12) | 0·96 (0·89 - 1·04) |
| 40-59 | 6,356 | 1,804 (28·4%) |  | reference | reference |
| 60-79 | 3,008 | 760 (25·3%) |  | 0·85 (0·77 - 0·94) | 0·96 (0·87 - 1·07) |
| ≥80 | 608 | 218 (35·9%) |  | 1·41 (1·18 - 1·68) | 1·59 (1·33 - 1·91) |
| **Gender** |  |  | < 0·001 |  |  |
| Female | 8,976 | 2,761 (30·8%) |  | reference | reference |
| Male | 7,888 | 2,082 (26·4 %) |  | 0·81 (0·75 - 0·86) | 0·79 (0·74 - 0·85) |
| **Healthcare Worker** |  |  | < 0·001 |  |  |
| No | 10,294 | 2,542 (24·7%) |  | reference | reference |
| Yes | 3,813 | 1,540 (40·4%) |  | 2·07 (1·91 - 2·24) | 2·77 (2·52 - 3·05) |
| **Outbreak Secondary Transmission** |  |  | < 0·001 |  |  |
| Index case | 13,904 | 3,483 (25·1%) |  | reference | reference |
| 1-5 days | 588 | 239 (40.6%) |  | 2.05 (1.73 - 2.43) | 2.22 (1.86 - 2.64) |
| 6-10 days | 592 | 265 (44.8%) |  | 2.42 (2.05 - 2.86) | 2.63 (2.21 - 3.12) |
| ≥11 days | 1,819 | 872 (47.9%) |  | 2.76 (2.49 - 3.04) | 2.68 (2.41 - 2.98) |
| **Household Secondary**<br>**Transmission** |  |  | < 0.001 |  |  |
| Index case | 14,124 | 3,882 (27.5%) |  | reference | reference |
| 1-5 days | 1,599 | 518 (32.4%) |  | 1.26 (1.13 - 1.41) | 1.12 (1.00 - 1.26) |
| 6-10 days | 690 | 245 (35.5%) |  | 1.45 (1.24 - 1.71) | 1.34 (1.14 - 1.58) |
| ≥11 days | 490 | 214 (43.7%) |  | 2.05 (1.70 - 2.45) | 1.73 (1.43 - 2.09) |
| **Visible Minority** | 34.9 (16.1, 65.6) | 36.2 (17.5, 65.6) | 0.010 | 1.02 (1.00 - 1.03) | 0.96 (0.94 - 0.97) |
| **Recent Immigration** | 3.9 (1.6, 7.5) | 4.2 (1.8, 7.5) | 0.001 | 1.13 (1.05 - 1.22) | 0.88 (0.80 - 0.98) |
| **Low-income Cut-off** | 11.5 (5.8, 17.9) | 12.1 (5.9, 18.3) | 0.128 | 1.03 (0.99 - 1.07) | 0.90 (0.86 - 0.95) |
| **Labour Force** | 63.7 (59.3, 68.7) | 64.0 (59.5, 69.0) | 0.010 | 1.07 (1.02 - 1.12) | 1.18 (1.09 - 1.22) |
| **No High School Education** | 18.1 (13.6, 23.6) | 18.4 (13.6, 23.9) | 0.298 | 1.02 (0.98 - 1.07) | 0.92 (0.88 - 0.96) |
| **Urban/Rural** |  |  | 0.334 |  |  |
| Rural | 2931 | 821 (28.0%) |  | reference | reference |
| Urban | 13,893 | 4,015 (28.9%) |  | 1.04 (0.96 - 1.14) | 0.84 (0.75 - 0.93) |
| **Region** |  |  | < 0.001 |  |  |
| Central East | 3,801 | 1,088 (28.6%) |  | 0.96 (0.85 - 1.09) | 0.84 (0.73 - 0.96) |
| Central West | 2,390 | 623 (26.1%) |  | 0.85 (0.74 - 0.97) | 0.76 (0.66 - 0.88) |
| East | 1,665 | 489 (29.4%) |  | reference | reference |
| North | 244 | 56 (23.0%) |  | 0.72 (0.52 - 0.98) | 0.84 (0.61 - 1.17) |
| South West | 1,660 | 413 (24.9%) |  | 0.80 (0.68 - 0.93) | 0.73 (0.62 - 0.86) |
| Toronto | 7,140 | 2,189 (30.7%) |  | 1.06 (0.95 - 1.20) | 0.84 (0.74 - 0.95) |

†Adjusted models included age, gender, month of symptom onset, and region.

**Table 3:**
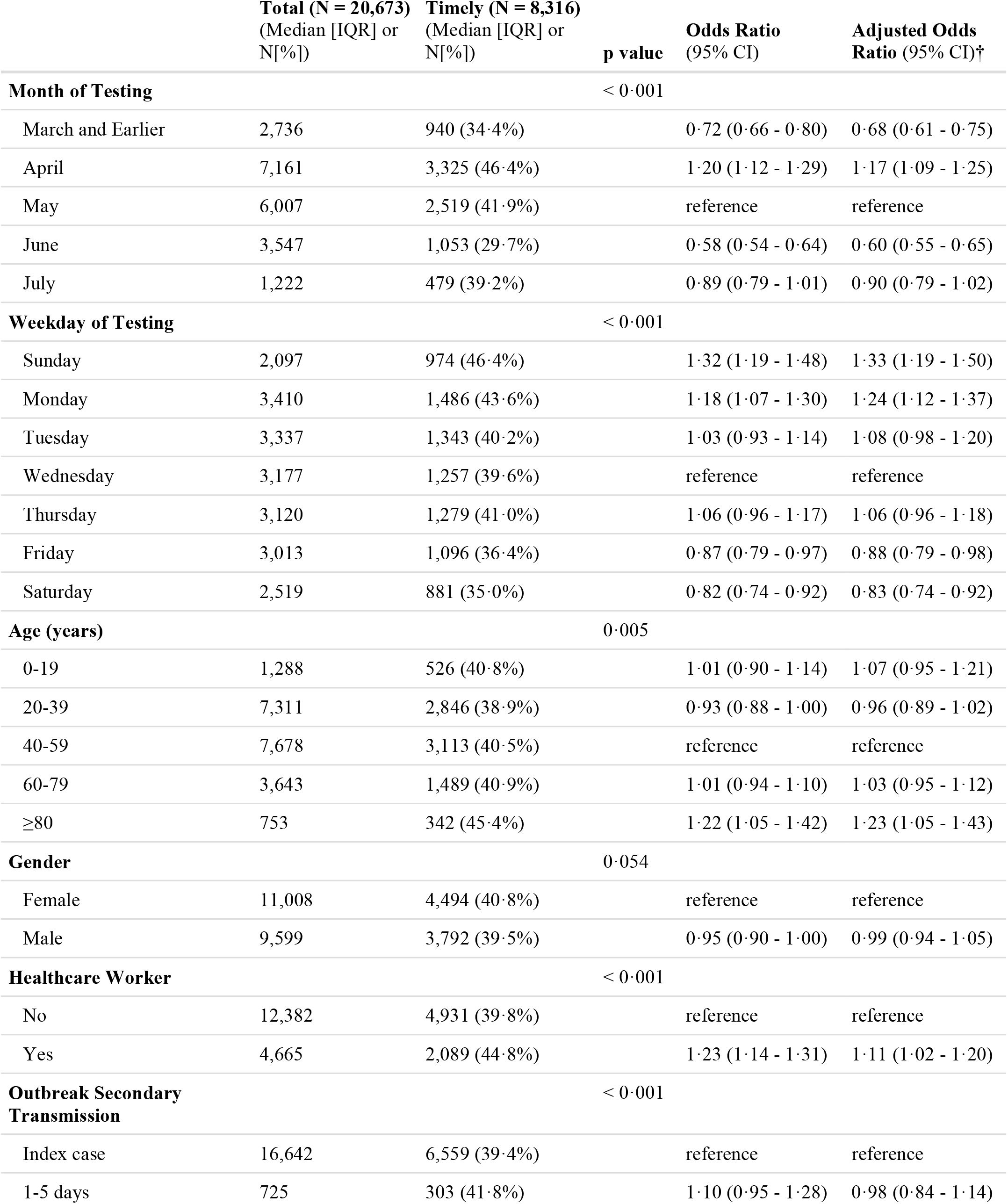
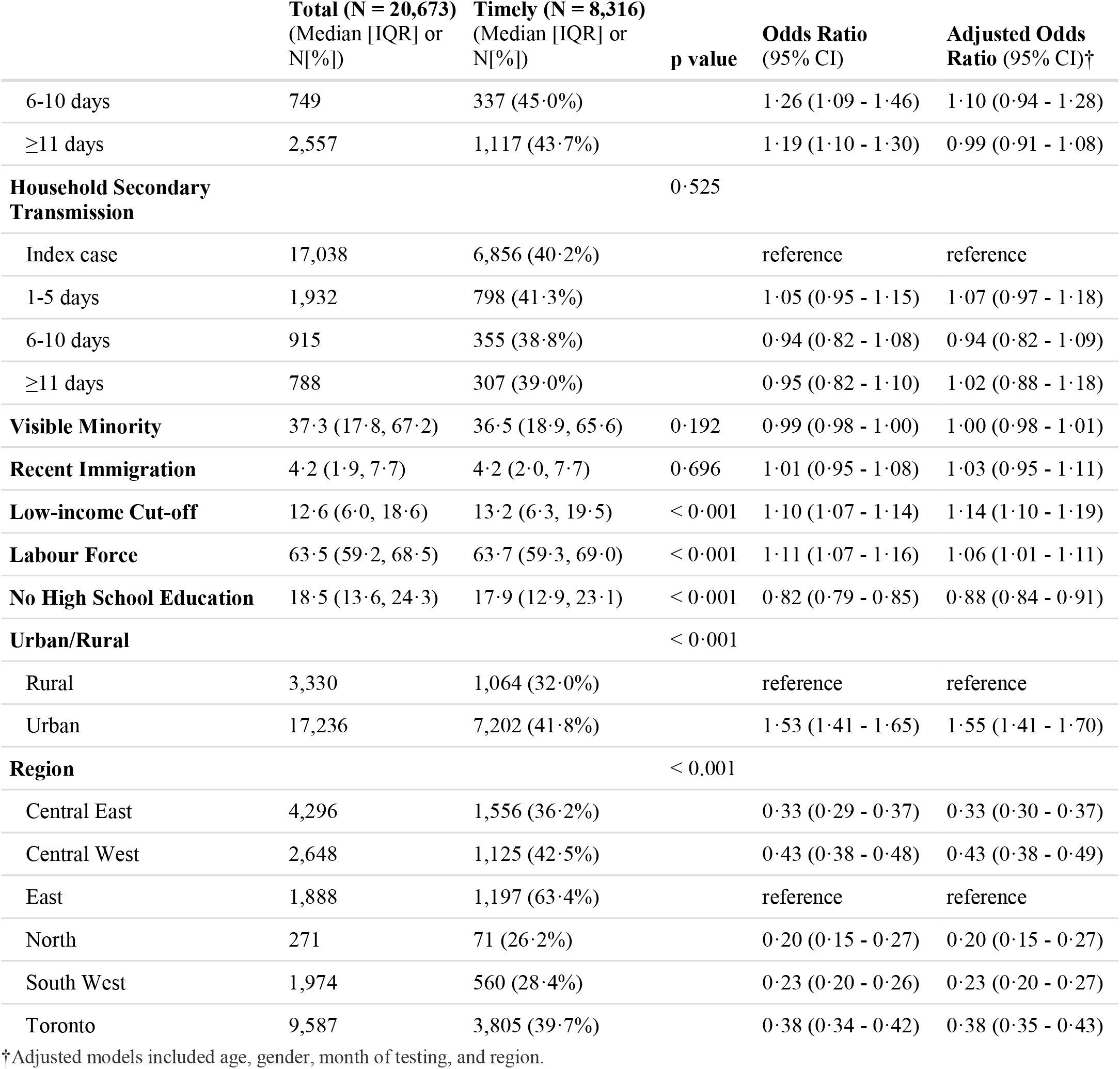
Factors associated with timely test turnaround.

### Timely Test Turnaround

Test turnaround for specimens collected on Fridays (aOR = 0·88 [95% CI: 0·79-0·98]) and Saturdays (aOR = 0·83 [95% CI: 0·74-0·92]; reference = Wednesday) were less likely to be timely. Of the individual-level characteristics, healthcare workers (aOR = 1·11 [95% CI: 1·02-1·20]) and those ≥80 years old (aOR = 1·23 [95% CI: 1·05-1·43]) had higher odds of timely test turnaround. Urban areas were more likely to have timely test turnaround compared to rural areas (aOR = 1·55 [95% CI: 1·41-1·70]).

### Timely Reporting

Temporal trends of increasing timely reporting were observed, where March had significantly lower odds of timely reporting (Table 4; aOR = 0·40 [95% CI: 0·36-0·44]) compared to May, whereas reporting in June (aOR = 5·25 [95% CI: 4·67-5·91]) and July (aOR = 8·90 [95% CI: 7·00-11·31]) was much more likely to be timely. Urban areas had lower odds of timely test reporting compared to rural areas (aOR = 0·79 [95% CI: 0·70-0·89]). Lower unadjusted odds of timely reporting were observed in neighbourhoods with a higher proportion of recent immigration (OR = 0·46 per 10% increase [95% CI: 0·43-0·49]), visible minority (OR = 0·91 per 10% increase [95% CI: 0·90-0·92]), and LICO (OR = 0·62 per 10% increase [95% CI: 0·60-0·64]). When adjusting age, gender, month of reporting and region, the associations weakened for recent immigration (aOR = 0·85 per 10% increase [95% CI: 0·79-0·93]), visible minority (aOR = 0·97 per 10% increase [95% CI: 0·95-0·98]), and LICO (aOR = 0·99 per 10% increase [95% CI: 0·94-1·04]). Age and secondary case status were not strongly associated with timely reporting after adjustment.

**Table 4:**
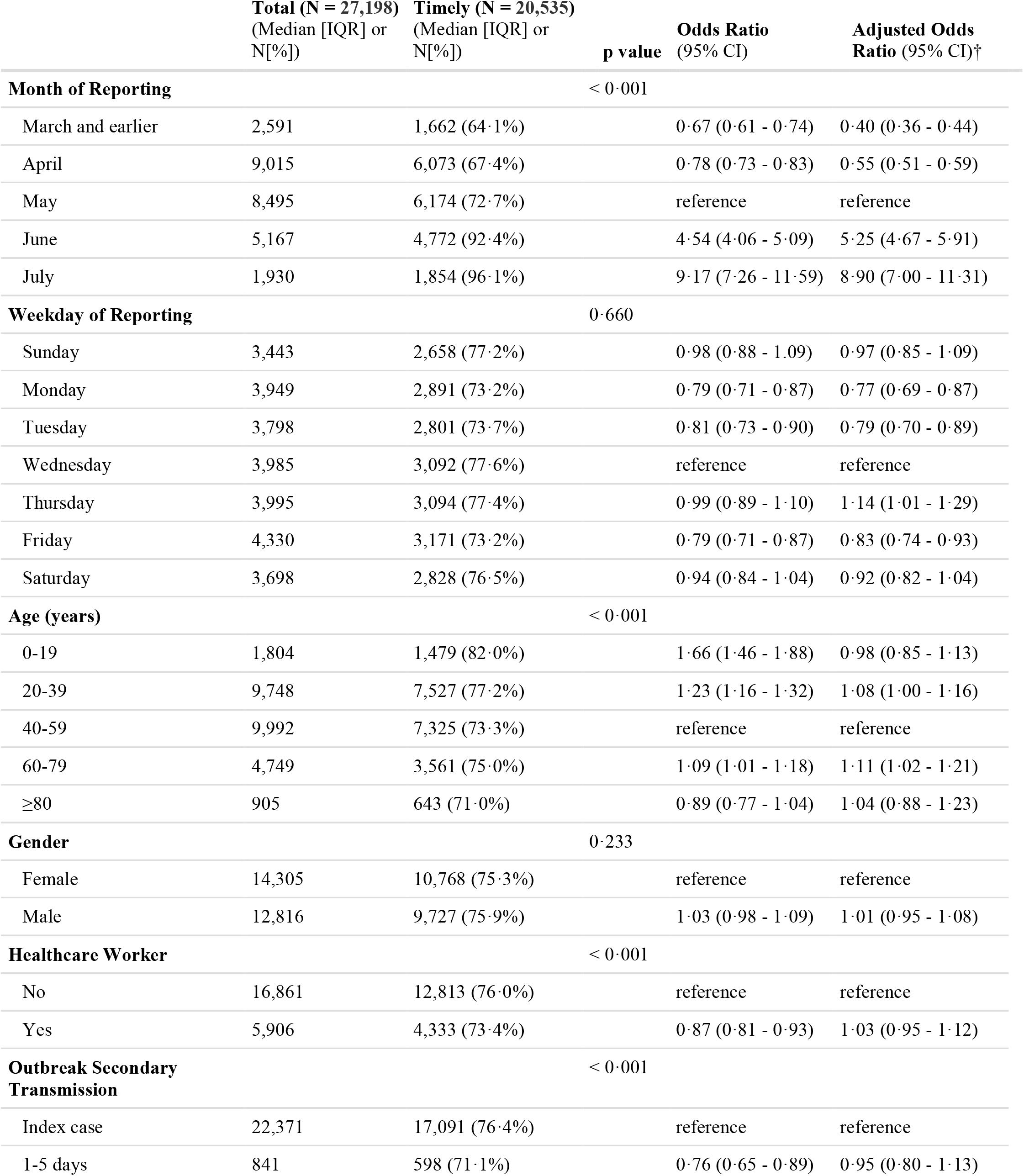
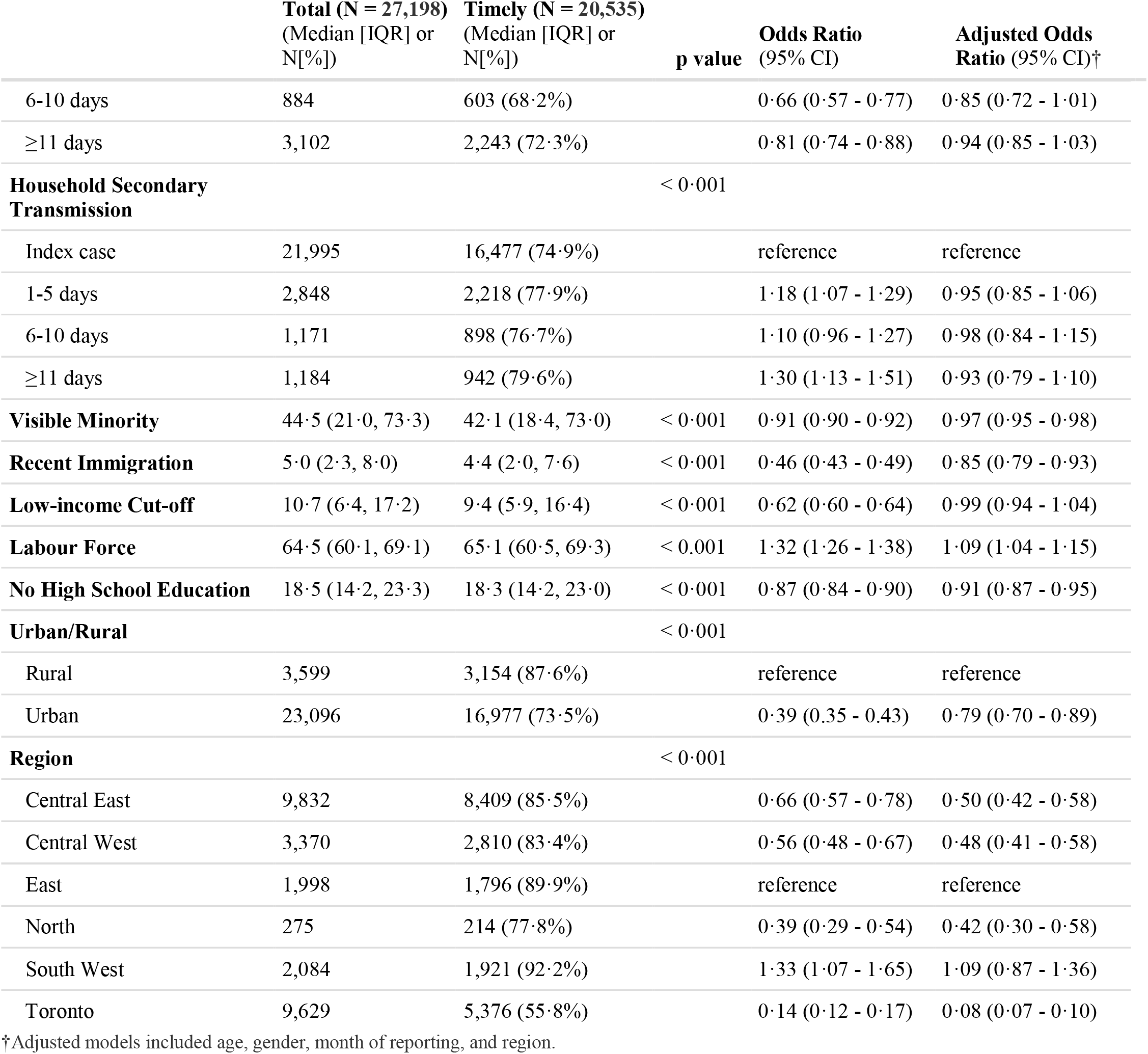
Factors associated with timely reporting.

## Discussion

In this study we examined delays in testing and reporting from a cohort of 27,198 confirmed COVID-19 cases from non-congregate settings in Ontario, and identified factors associated with timely test seeking, test turnaround, and reporting. Factors associated with timely test seeking included female gender, healthcare workers, individuals aged ≥80 years, and a previously identified index case within the same household; factors associated with timely test turnaround included age ≥80 years, healthcare worker status, and residence in an urban area; and factors associated with timely reporting included month of reporting and residence in a rural areas. We noticed important weekend effects wherein there was decreased timeliness for test seeking, test turnaround, and reporting on Fridays and Saturdays.

Our analysis identified that test seeking was frequently delayed in our cohort. Various mathematical modelling studies have highlighted the importance of shortening the delay between symptom onset to diagnosis for effectively controlling COVID-19 outbreaks.^10^ Kretzschmar et al. found testing delays strongly influenced the ability of contact tracing strategies to reduce the effective reproduction number (Re), such that testing delays of 3 days or longer preclude reduction of Re below 1, even with 100% tracing coverage.^9^ Hellewell et al. identified symptom onset to test delays play a major role in the probability of controlling an outbreak.^6^ Rong et al. conducted a study based on reported data from China in the first 3 months of the COVID-19 pandemic, and identified that the proportion of timely diagnosis was important in reducing the number of new infections and further transmission risk.^12^ Model results from Larremore et al. highlighted the increased importance of speed of testing, reporting, and contact tracing, over improved test sensitivities, to control COVID-19.^13^ Additional studies have identified the importance of faster testing from symptom onset to lower the risk of household transmission.^7,14,15^ While these studies suggest that increased timeliness of test seeking can lead to decreases in COVID-19 transmission, our study uses empirical evidence to identify factors associated with timeliness.

The trends of increasing timely test seeking during the study period were likely driven by changes in testing criteria and test access.^16^ Healthcare workers and vulnerable populations were recommended by the World Health Organization early in the pandemic for testing prioritization.^17^ In April, the provincial government of Ontario expanded testing to include these groups, specifically individuals working in healthcare settings.^18^ On May 29 testing in Ontario was opened to all individuals.^19^ The expanded testing criteria and increased test access via the rollout of assessment centre testing likely contributed to the increased timely test seeking observed in June and July. The gender differences for test seeking observed in this cohort may be explained by patterns of delayed healthcare seeking behaviour among men.^20^ Secondary cases of index cases linked to an outbreak or within a household were more likely to have timely test seeking than the index case. Our analysis describes the effect of testing criteria and factors associated with timely test seeking, which may be used to inform strategies such as expanded hours for testing including weekends, increased access to testing for working populations.

Timely test turnaround was more likely in healthcare workers compared to non-healthcare workers. A cross-sectional study of healthcare workers in US nursing homes identified 13·5% of facilities had test turnaround times within 1 day, and 16·4% in hot-spot counties.^21^ We observed a higher proportion of timely test turnaround in our cohort, as this may be more reflective of different strategies and testing resources available across jurisdictions. Urban areas were more likely to have timely test turnaround compared to rural ones. This likely reflects the larger laboratory capacity in urban centers and challenges in the transport of samples in more rural areas. The reduction of timely test turnaround in June was likely driven by a reporting error a local hospital network that impacted roughly 500 cases.^22^ Our analysis also found timely test turnaround was less likely on Fridays and Saturdays. Weekend effects have been described for other facets of medical care,^23^ but this has not been previously described in medical laboratory testing. These weekend effects may be influenced by lower frequency in sample transportation that contributes to longer delays in test turnaround. This effect on timely testing may be determined by changes in laboratory capacity and staffing over the weekend.

Increasingly rapid reporting was observed from March to July and was likely driven by increasing local public health capacity in response to the growth of COVID-19 in Ontario and directives issued to local public health for required timely data entry of case information. Urban areas and regions that experienced large numbers COVID-19 cases in their jurisdictions, such as Toronto, were less likely to have timely reporting to the provincial database. This may also explain the lower odds of timely reporting observed for neighbourhoods with higher proportions of recent immigrants, visible minority, and low income as these neighbourhoods were typically found around the Toronto region. This highlights the impact of exponential growth and large outbreaks that may rapidly surge beyond public health capacity causing delays in timely disease reporting precisely when they are needed most.

Our study has limitations as this cohort was generated from cases arising from the first wave of COVID-19 in Ontario, and these associations may not be generalizable to other populations/jurisdictions or subsequent time periods. There may be a degree of misclassification with index and secondary cases within households if members were not tested or if secondary cases had been infected outside of the home. Another limitation is that our outcomes of interest had incomplete data, specifically the test seeking and test turnaround delays. While the delay from symptom onset is likely a reasonable proxy of infection onset to test seeking for symptomatic patients, the delay in test seeking could not be measured relative to the start of true infectiousness among asymptomatic infections. Delays in timely test seeking were likely influenced by changes in test access, testing criteria, and laboratory capacity during our study period. Further, missing information on specimen collection led to incomplete test seeking and test turnaround outcomes. The large reporting error in June likely resulted in decreased timely test turnaround in our analysis. Changes in required data entry fields over our study period may have also impacted timely reporting. However the strengths of our study include a large population-based cohort; an examination of multiple delays in the disease reporting process; and a large number of individual, structural, and neighbourhood characteristics, including the identification of index and secondary cases within households. Our study provides empirical evidence on factors associated timely disease reporting, whereas many prior studies focused on modelling delays. To our knowledge this is the first study describing detailed delays in COVID-19 reporting in Canada.

This analysis described the delays in test seeking, test turnaround, and reporting across a cohort of community COVID-19 cases in Ontario. We identified numerous factors associated with timeliness which could be used to inform interventions. In particular, weekend effects were apparent for all delays, suggesting that expanded awareness, access, and capacity over the weekends may reduce delays. We also found improvements in timeliness for each stage of the disease reporting process over the course of the first pandemic wave. The findings of this study may be used to develop strategies to improve timely testing and reporting of SARS-CoV-2 infections.

## Data Availability

Data sharing requests should be directed to Public Health Ontario.

## Contributors

EJ performed the analysis and drafted the manuscript. KB, SB, and ND conceptualized the study. KB, SB, ND, and EJ developed the methodology. KB, SB, and EJ verified the underlying data. LP contributed to the analysis. KB, SB, ND, and LP reviewed the manuscript.

## Declaration of interests

The authors have no conflicts of interest to declare.

## Data sharing

Data sharing requests should be directed to Public Health Ontario.

## Notes

### Competing Interest Statement

The authors have declared no competing interest.

### Author Declarations

We obtained ethics approval from Public Health Ontario's Research Ethics Board.

