## Supplementary Material for "Factors Associated with Timely Test Seeking, Test Turnaround, and Public Reporting of COVID-19: a retrospective analysis in Ontario, Canada"

| | |
|---|---|
| Eugene Joh, MPH[a] | |
| Sarah A. Buchan, PhD[a–c] | |
| Nick Daneman, MD[a,d–f] | |
| Lauren A. Paul, MSc[a] | |
| Kevin A. Brown, PhD[a–c] | |

*Affiliations:*

[a]Health Protection, Public Health Ontario, 661 University Ave., Floor 17, Toronto, ON, M5G 1M1, Canada

[b]ICES, 2075 Bayview Ave., Room G1 06, Toronto, ON, M4N 3M5, Canada

[c]Dalla Lana School of Public Health, University of Toronto, 155 College St., Room 500, Toronto, ON, M5T 3M7, Canada

[d]Sunnybrook Research Institute, Sunnybrook Health Sciences Centre, 2075 Bayview Ave., Toronto, ON, M4N 3M5, Canada

[e]Division of Infectious Diseases, Sunnybrook Health Sciences Centre, 2075 Bayview Ave., Room B1 03, Toronto, ON, M4N 3M5, Canada

[f]Institute of Health Policy, Management and Evaluation, University of Toronto, 155 College St., Suite 425, Toronto, ON, M5T 3M6, Canada

*Corresponding Author:*

Eugene Joh, MPH
Public Health Ontario
661 University Avenue, Floor 17
Toronto, Ontario, M5G 1M1


### Supplementary Tables

**Supplementary Table S1.** Health regions and public health units in Ontario, Canada

| Health Region | Public Health Units |
|---|---|
| North | Algoma Public Health |
| | North Bay Parry Sound District Health Unit |
| | Northwestern Health Unit |
| | Porcupine Health Unit |
| | Public Health Sudbury & Districts |
| | Thunder Bay District Health Unit |
| | Timiskaming Health Unit |
| East | Eastern Ontario Health Unit |
| | Hastings Prince Edward Public Health |
| | Kingston, Frontenac and Lennox & Addington Public Health |
| | Leeds, Grenville & Lanark District Health Unit |
| | Ottawa Public Health |
| | Renfrew County and District Health Unit |
| Central East | Durham Region Health Department |
| | Haliburton, Kawartha, Pine Ridge District Health Unit |
| | Peel Public Health |
| | Peterborough Public Health |
| | Simcoe Muskoka District Health Unit |
| | York Region Public Health |
| Central West | Brant County Health Unit |
| | City of Hamilton Public Health Services |
| | Haldimand-Norfolk Health Unit |
| | Halton Region Public Health |
| | Niagara Region Public Health |
| | Region of Waterloo Public Health and Emergency Services |
| | Wellington-Dufferin-Guelph Public Health |
| Toronto | Toronto Public Health |
| South West | Chatham-Kent Public Health |
| | Grey Bruce Health Unit |
| | Huron Perth Public Health |
| | Lambton Public Health |
| | Middlesex-London Health Unit |
| | Southwestern Public Health |
| | Windsor-Essex County Health Unit |
